# Cardiorespiratory training after stroke or transient ischemic attack in the United Kingdom: a national survey

**DOI:** 10.1101/2025.11.01.25339298

**Authors:** P Hartley, K Bond, H Probert, C Brown, K Khadjooi, J McPeake

**Affiliations:** Department of Physiotherapy, Cambridge University Hospitals NHS Foundation Trust, Cambridge, UK; The Healthcare Improvement Studies Institute, University of Cambridge, Cambridge, UK; Department of Cardiovascular Prevention and Rehabilitation, Guy’s and St Thomas’ NHS Foundation Trust, London, UK; Department of Stroke Medicine, Cambridge University Hospitals NHS Foundation Trust, Cambridge, UK; School of Clinical Medicine, University of Cambridge, Cambridge, UK

## Abstract

**Objective:** To assess the current provision and beliefs regarding benefits and risks, barriers, and potential future service models of cardiorespiratory training after stroke and transient ischaemic attack (TIA) in the United Kingdom.

**Methods:** An online survey of physiotherapists and exercise professionals working in stroke or cardiac rehabilitation.

**Results:** The 253 respondents were separated into groups, Group 1: those integrating cardiorespiratory training into routine stroke rehabilitation (n=113); Group 2: those in specialist cardiorespiratory services including people after stroke or TIA (n=40); Group 3: those in stroke rehabilitation not integrating cardiorespiratory training (n=74); Group 4: those in specialist cardiorespiratory services not seeing people after stroke or TIA (n=26).

Delivery of cardiorespiratory training varied in frequency and duration of supervised sessions, and people with minimal disability may be more likely to receive cardiorespiratory training.

Identified stroke-specific risk factors for cardiorespiratory training varied both in the factors identified, and whether they were considered precautions or contraindications.

The most frequently identified concerns were risk of serious adverse events, fatigue, and falls. The most common barriers to delivering cardiorespiratory training included commissioning of services, resources, skills and knowledge, and a lack of guidelines.

**Conclusions:** Most respondents reported providing cardiorespiratory training after stroke or TIA, though many people who have a stroke or TIA may not receive this training. The results highlight a potential inequality where people with greater disability may be less likely to receive or be eligible for cardiorespiratory training. There are many factors making cardiorespiratory training a complex challenge to deliver in clinical practice.

## Background

Strokes and transient ischaemic attacks (TIAs) have lifechanging effects on individuals [1-3] and families [4], and significantly impact health and social care systems [5]. The 5-year risk of recurrent stroke after a TIA or stroke is 9.5-12%.[6, 7], and the 5-year risk of acute coronary syndrome after ischaemic stroke is 11.1% [8].

Cardiorespiratory training after stroke can improve cardiorespiratory fitness and disability [9-12], and evidence on its impact on cardiovascular risk factors is promising, though inconclusive [9]. As such, mirroring international guidelines [13-15], regarding long-term management and secondary prevention, the 2023 National Clinical Guideline for Stroke for the United Kingdom (UK) and Ireland recommends that: “people with stroke should be offered cardiorespiratory training or mixed training regardless of age, time since having the stroke, and severity of impairment”[16].

Whilst there are known to be many factors influencing effective implementation of cardiorespiratory training in rehabilitation after stroke, evidence pertaining to the UK is very limited [17]. Therefore, the aim of the project is to assess the current provision and beliefs regarding benefits and risks, barriers, and potential future service models of cardiorespiratory training after stroke and TIA in the UK from the perspective of physiotherapists and exercise professionals.

## Methods

The survey was conducted online using Thiscovery (https://www.thiscovery.org), an online research and collaboration platform. In the absence of a validated questionnaire, the survey was created by the research team using the available literature [17-21] and with input from clinicians. The survey was pilot tested by five clinicians and refined based on their feedback.

Participants were physiotherapists or exercise professionals whose work includes stroke or cardiac rehabilitation in the UK. They were recruited via social media, and various professional networks including, the Association of Chartered Physiotherapists in Neurology, the Association of Chartered Physiotherapists in Cardiovascular Rehabilitation, the British Association for Cardiovascular Prevention and Rehabilitation, the Integrated Stroke Delivery Network, Chartered Society of Sport and Exercise Scientists (CASES), and the Scottish Stroke AHP Forum. Potentially interested participants followed a web link to a project homepage which included a participant information sheet. Before starting the survey participants had to provide electronic informed consent.

The survey filtered respondents to different questions based on the service they worked in and whether they currently delivered cardiorespiratory training to people after stroke or TIA. The survey sections included: current practice of cardiorespiratory training after stroke or TIA; barriers to delivering cardiorespiratory training after stroke or TIA; their beliefs regarding the challenges and risks of cardiorespiratory training after stroke or TIA; and a brief section on demographics. The full survey is available in the online supplementary materials.

The study protocol was given a favourable opinion by the Cambridge Psychology Research Ethics Committee, University of Cambridge.

The quantitative analysis was conducted using R (version 4.3.2) [22]. Descriptive statistics are presented as count and percentage, or as mean and standard deviation, or median and interquartile range if non-normally distributed. Free-text responses were analysed using content analysis. One member of the research team coded the free-text (one free-text response could be assigned to multiple categories); the naming of categories, and coding of data were confirmed by a second researcher, and summarised quantitatively. Respondents were separated into four groups, those that incorporated cardiorespiratory training within routine stroke rehabilitation; those that provide cardiorespiratory training within specialist services (such as cardiac rehabilitation) to people with stroke or TIA; those that work within stroke rehabilitation but do not (or rarely) provide cardiorespiratory training, and those that work in cardiac rehabilitation or similar services but do not (or rarely) provide cardiorespiratory training.

For categorical variables, if two groups answered the same questions, the proportions were analysed with a chi-squared test of independence or if an expected cell count was <5 we used Fisher’s exact test. For categorical variables answered by all four groups we used Fisher’s exact test on the four-by-two contingency table. If significant differences were found, post-hoc pairwise comparisons of proportions with Yates’ continuity correction and Holm-adjusted p-values were performed.

For the continuous variables, differences between two groups were assessed with Student’s t-test if data normally distributed, and Mann-Whitney U test if not. Comparisons between four groups of variables were assessed with a Kruskal–Wallis test.

## Results

A total of 255 people consented to the survey and were eligible to take part, however 2 people did not complete any questions leaving 253 respondents. Of the 253 participants, 207 completed the survey providing a completion rate of 82% (Figure 1). Table 1 describes the characteristics of the respondents.

**Table 1.** Participant characteristics.

|  | Group 1: Integrating CR into stroke rehabilitation (n = 113) | Group 2: CR training service including stroke (n = 40) | Group 3: Not integrating CR into stroke rehabilitation (n = 74) | Group 4: CR training service not including stroke (n = 26) |
| --- | --- | --- | --- | --- |
| **Profession** |  |  |  |  |
| Physiotherapist | 82 (72.57%) | 14 (35.00%) | 58 (78.38%) | 11 (42.31%) |
| Clinical exercise physiologist | 2 (1.77%) | 6 (15.00%) | 0 (0.00%) | 3 (11.54%) |
| Other exercise professional | 7 (6.19%) | 10 (25.00%) | 4 (5.41%) | 5 (19.23%) |
| Prefer not to say | 1 (0.88%) | 0 (0.00%) | 1 (1.35%) | 2 (7.69%) |
| **Years of Professional Practice (median IQR)** | 15.00 (6.50-22.00) | 18.00 (11.00-23.50) | 16.50 (7.25-22.00) | 16.00 (10.50-24.50) |
| **Region** |  |  |  |  |
| Scotland | 3 (2.65%) | 3 (7.50%) | 9 (12.16%) | 1 (3.85%) |
| Wales | 1 (0.88%) | 1 (2.50%) | 0 (0.00%) | 1 (3.85%) |
| Northern Ireland | 0 (0.00%) | 0 (0.00%) | 0 (0.00%) | 2 (7.69%) |
| England, London | 17 (15.04%) | 2 (5.00%) | 7 (9.46%) | 3 (11.54%) |
| England, East of England | 21 (18.58%) | 7 (17.50%) | 10 (13.51%) | 5 (19.23%) |
| England, South East | 19 (16.81%) | 4 (10.00%) | 10 (13.51%) | 5 (19.23%) |
| England, South West | 6 (5.31%) | 3 (7.50%) | 9 (12.16%) | 2 (7.69%) |
| England, North West | 5 (4.42%) | 5 (12.50%) | 3 (4.05%) | 0 (0.00%) |
| England, North East | 2 (1.77%) | 0 (0.00%) | 1 (1.35%) | 1 (3.85%) |
| England, East Midlands | 7 (6.19%) | 2 (5.00%) | 2 (2.70%) | 0 (0.00%) |
| England, West Midlands | 4 (3.54%) | 2 (5.00%) | 6 (8.11%) | 0 (0.00%) |
| England, Yorkshire and the Humber | 8 (7.08%) | 1 (2.50%) | 6 (8.11%) | 1 (3.85%) |
| **Sector** |  |  |  |  |
| NHS | 101 (89.38%) | 30 (75.00%) | 68 (91.89%) | 21 (80.77%) |
| Private practice | 8 (7.08%) | 3 (7.50%) | 3 (4.05%) | 0 (0.00%) |
| Charitable | 2 (1.77%) | 4 (10.00%) | 1 (1.35%) | 0 (0.00%) |
| Local authority | 1 (0.88%) | 2 (5.00%) | 1 (1.35%) | 3 (11.54%) |
| Other | 1 (0.88%) | 1 (2.50%) | 1 (1.35%) | 2 (7.69%) |
| **Service** |  |  |  |  |
| Inpatient stroke/rehabilitation unit | 45 (39.82%) | 0 (0.00%) | 26 (35.14%) | 1 (3.85%) |
| Community neuro-rehabilitation service | 53 (46.90%) | 2 (5.00%) | 37 (50.00%) | 1 (3.85%) |
| Community rehabilitation team linked with short-term care service | 8 (7.08%) | 0 (0.00%) | 2 (2.70%) | 0 (0.00%) |
| Inpatient and community stroke rehabilitation | 0 (0.00%) | 0 (0.00%) | 4 (5.41%) | 0 (0.00%) |
| Cardiac Rehabilitation | 2 (1.77%) | 30 (75.00%) | 1 (1.35%) | 22 (84.62%) |
| Inpatient cardiology wards | 0 (0.00%) | 2 (5.00%) | 0 (0.00%) | 0 (0.00%) |
| Other outpatient/community cardiac service | 0 (0.00%) | 1 (2.50%) | 0 (0.00%) | 2 (7.69%) |
| Outpatient/community respiratory service | 0 (0.00%) | 1 (2.50%) | 0 (0.00%) | 0 (0.00%) |
| Other community exercise programmes | 0 (0.00%) | 4 (10.00%) | 0 (0.00%) | 0 (0.00%) |

**Figure 1.**
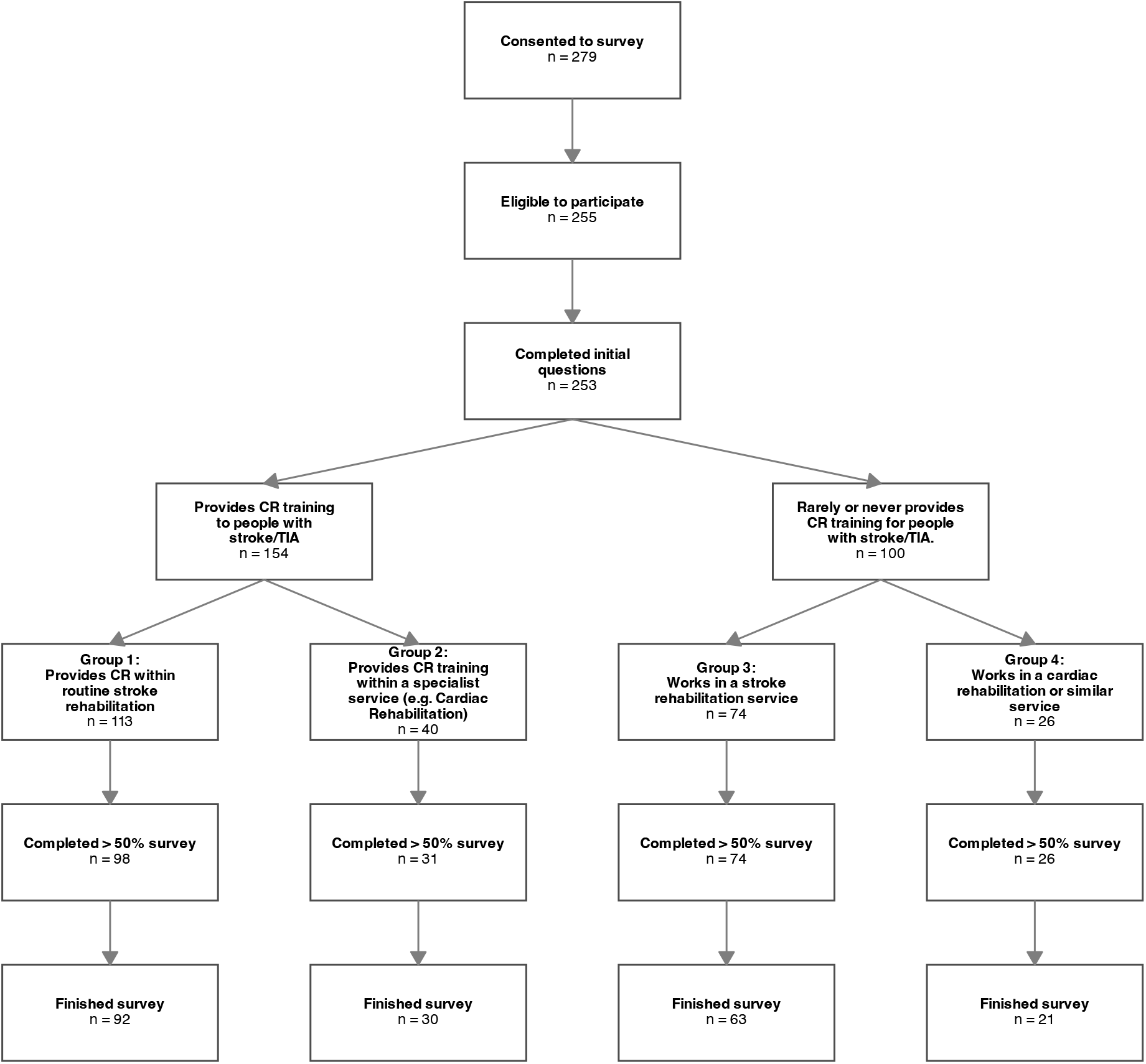
Participant flow through survey

### Current provision of cardiorespiratory training after stroke and TIA

A total of 153 participants (60%) reported providing cardiorespiratory training to people after stroke and TIA. This includes 113 respondents who reported incorporating cardiorespiratory training into routine stroke rehabilitation (Group 1) and 40 respondents who reported working in specialist services that deliver cardiorespiratory training to people, including those who have had a stroke or TIA (Group 2). In this later group of respondents, the median estimated proportion of service users who have a diagnosis of stroke was 5% (IQR: 3 to 5%), and for TIA, also 5% (IQR: 3 to 10%).

Participants in Groups 1 and 2 were asked what the typical frequency, and duration in weeks of their supervised cardiorespiratory programmes were. The most common frequency of supervised sessions reported was once per week (online supplementary materials table A). Answered regarding duration ranged from 1 week to unlimited durations, and 11 participants reported that duration was too variable to estimate or was dependent on length of stay in hospital or NHS rehabilitation. In those that gave a figure (where respondents gave a range e.g. 4 to 6 weeks, the midpoint was taken), the median was 8.0 weeks (IQR: 6.0-10.0). There was no significant differences in duration between those integrating cardiorespiratory training in routine stroke rehabilitation (Group 1) and those in specialist cardiorespiratory services (Group 2), (p = 0.868).

Respondents integrating cardiorespiratory training in routine stroke rehabilitation (Group 1) and those in specialist cardiorespiratory services (Group 2) were asked about the typical time after stroke or TIA that they would provide the training. The most frequently selected option in the group integrating the training into routine cardiorespiratory training (Group 1) was 3-12 weeks after stroke (42%), although there was considerable variation. Whereas the most frequently selected option in the respondents working within a specialist cardiorespiratory service (Group 2) was 13 or more weeks (39.4%). Whilst there were variations in both groups, there was a significantly greater proportion of respondents selecting 0-2 weeks in the routine stroke rehabilitation group (23% vs 3%, p = 0.02) and a significantly greater proportion of respondents selecting 13+ weeks in the respondents working in specialist cardiorespiratory service (17% vs 39%, p = 0.01).

Respondents who reported that they provide cardiorespiratory training as part of routine stroke rehabilitation (Group 1) were asked which patients they were more likely to incorporate cardiorespiratory training with (online supplementary materials table B). Of these respondents 41 (38%) reported incorporating cardiorespiratory training with all their patients. The only multiple-choice answers to be selected by over 50% of respondents were individuals with minimal physical disability (55%); individuals with minimal cognitive disability (53%); and individuals who were more active before their stroke or TIA (51%). It is worth noting that there were contrary responses to the most common answers, such as 19% of respondents selecting that they would be more likely to incorporate cardiorespiratory training with individuals with significant physical disability.

Respondents who reported that they provide training to people with stroke or TIA as part of a specialist service delivering cardiorespiratory training (Group 2) were asked how people who have had a stroke or TIA are eligible for their service (online supplementary materials table C). The majority (74%) reported that people who have a diagnosis of stroke would only be eligible if they have a secondary diagnosis such as a cardiac condition, with a similar proportion reporting the same for a diagnosis of TIA (68%). Survey participants were also asked whether people with stroke are eligible regardless of the severity of their stroke impairments and disability (online supplementary materials table D). Half reported that there were no eligibility criteria that relate to the severity of stroke impairments or disability. However, 29% reported that participants must be able to exercise safely unsupervised, and 26% reported eligibility criteria relating to physical impairments, and 18% criteria relating to cognitive impairments.

### Risks and concerns with cardiorespiratory training after stroke and TIA

Both Group 1 and Group 2 were asked to select stroke-specific precautions or contraindications to cardiorespiratory training that they use, with the option to add free text (online supplementary materials table E and F). Only cervical arterial dissection was selected by at least half (51%) of all respondents as either a precaution (59% of those selecting cervical arterial dissection) or contraindication (41% of those selecting cervical arterial dissection).

In a separate question all respondents (Groups 1 to 4), including those that do not (or only rarely) deliver cardiorespiratory training after stroke or TIA were asked what concerns (if any) they have about incorporating cardiorespiratory training into the rehabilitation of people who have had a stroke or TIA (online supplementary materials table G). Whilst 14% reported no concerns, others reported a number of concerns. The most frequently identified concerns were risk of serious adverse event whilst exercising such as a cardiovascular event (37%); risk of falling (25.3%); increased fatigue (27.7%); and an increase in compensatory or abnormal movement patterns (26.5%). Only risk of serious adverse event whilst exercising showed significant between-group differences in the proportion of respondents in each group identifying it as a concern (Group 1: 48%, Group 2: 18%, Group 3: 40.5%, Group 4: 8%, p = 0.002). Post-hoc testing showed significantly higher proportions of respondents concerned by the risk of serious adverse events in the group integrating cardiorespiratory training into routine rehabilitation (Group 1) than either the groups working in specialist cardiorespiratory services (Group 1 vs Group 2: p <0.05; and Group 1 vs.

Group 4: p <0.05). Post-hoc testing also showed significantly higher proportion of respondents concerned by the serious adverse events in the group working in stroke rehabilitation but not (or rarely) integrating cardiorespiratory training (Group 3) compared to the group working in a specialist cardiorespiratory service, but not (or rarely) to people with stroke or TIA (Group 4) (p<0.05).

### Factors preventing delivery of cardiorespiratory training after stroke and TIA

We asked those not currently delivering cardiorespiratory training to people who have had a stroke or TIA (Groups 3 and 4) what was currently preventing them from doing so. Multiple barriers were identified, the most common of which included skills and knowledge regarding cardiorespiratory training (56%, though significantly higher proportions in stroke services group 73% vs 8%, p<0.05), and inadequate staffing (43%), and lack of guidelines regarding screening or risk-assessment (40%) (online supplementary materials table H).

### Confidence with delivery of cardiorespiratory training after stroke and TIA

Finally, we also asked respondents who were delivering cardiorespiratory training to people after stroke or TIA (Groups 1 and 2) to rate their current confidence to deliver cardiorespiratory training to people with stroke or TIA regardless of level of impairment from 1 (not at all confident) to 5 (fully confident). Those working in specialist services that deliver cardiorespiratory training had significantly higher confidence (median 4, IQR: 3 to 5) than those in stroke rehabilitation services (median 3, IQR: 3 to 5). Participants were asked if they were not fully confident, what would increase their confidence, and to provide a free-text response. The most common answers referred to training or education, and knowledge (42%), and second, referred to guidelines, including practical guidance for screening, monitoring and delivering (11%).

## Discussion

This survey highlighted large variations in cardiorespiratory delivery by physiotherapists and exercise professionals to people after stroke or TIA. Our results indicate that people with greater disability may be less likely to receive or be eligible for cardiorespiratory training. We also found large variations in recognised precautions and contraindications to cardiorespiratory training, as well as concerns for delivering cardiorespiratory training to this population highlighting potential clinical uncertainty. Training and education, as well as the availability of guidelines were identified both as barriers to delivery, as well as factors that would improve confidence in delivery of cardiorespiratory training after stroke.

That the majority of respondents reported delivering cardiorespiratory training either as part of routine stroke rehabilitation (Group 1) or in specialist cardiorespiratory services (Group 2) after stroke or TIA is positive. Although it is possible this positive finding is in part either the result of self-selection bias or even a social desirability bias given the recent guideline recommendations [16]. However, even amongst this majority, there appears to be more work to be done to be in line with the 2023 National Clinical Guideline for Stroke for the UK and Ireland guidelines [16]. First, the guidelines recommend that regardless of severity of impairment people with stroke should be offered cardiorespiratory training [16]. Many of those working in stroke rehabilitation services acknowledged they were more likely to incorporate cardiorespiratory training with those with minimal physical or cognitive impairment, and many of the specialist cardiorespiratory training had eligibility criteria that may exclude those with greater impairment. Second, the guidelines recommend a frequency of three to five times a week and a duration of 10-20 weeks [16]. Whilst we only asked about supervised exercise sessions, our data would suggest that sessions are rarely this frequent, and usually not meeting this duration.

Our findings regarding barriers to and concerns with delivering cardiorespiratory training after stroke are consistent with the findings of the review by Gaskins *et al*. [17]. Some of the factors identified in this survey such as the need for training and people not being referred to cardiorespiratory training services may be easily resolved compared to the commissioning of services and lack of resources. A current barrier to the commissioning of services and lack of resources may be the inconclusive evidence regarding the effect of cardiorespiratory training on improving cardiovascular risk factors after stroke [9], however, a recent US modelling study concluded that cardiac rehabilitation after stroke is cost-effective compared with usual care [23]. Hopefully these findings of cost-effectiveness will support further implementation of the aforementioned guideline recommendations.

The need for practical guidelines was identified by a number of respondents. Whilst there are extensive guidelines for cardiorespiratory training for people with heart disease including from the British Association for Cardiovascular Prevention and Rehabilitation [24], there is less guidance specific to stroke, although international recommendations do exist [13, 15]. As reflected in the results of this survey there remains uncertainty regarding the risks associated with more stroke specific risk factors such as atrial septal defects or cervical artery dissection, and exercise [25, 26].

Despite our best efforts, we had very limited representation from certain regions, including Wales and Northern Ireland. Generalisability would also have been greatly improved by further representation of exercise professionals and those working in specialist cardiorespiratory training services. As such, the findings are not necessarily generalisable to the whole of the UK.

The survey highlights that many physiotherapists and exercise professionals are delivering cardiorespiratory training to people after stroke and TIA, though there is significant variation. Even in those currently delivering cardiorespiratory training the data suggests that many are not currently implementing the 2023 National Clinical Guideline for Stroke for the UK and Ireland guidelines in full, including the inclusivity of those being provided with cardiorespiratory training, and frequency and dose of cardiorespiratory training. Guidelines focussing on the practical implementation of cardiorespiratory training after stroke would be welcomed by many of the survey respondents. These findings highlight the need for research to support additional resources and commissioning of services, including research investigating the cost-effectiveness of cardiorespiratory training after stroke and TIA, and its effect on cardiovascular risk factors and secondary stroke prevention.

## Data Availability

Anonymised data may be made available upon reasonable request.

## References

1. Douiri A, Rudd AG, Wolfe CD. Prevalence of poststroke cognitive impairment: South London Stroke Register 1995-2010. Stroke. 2013 Jan;44(1):138–45.

2. Marshall IJ, Wolfe C, Emmett E, Wafa H, Wang Y, Douiri A, Bhalla A, et al. Cohort profile: The South London Stroke Register - a population-based register measuring the incidence and outcomes of stroke. J Stroke Cerebrovasc Dis. 2023 Aug;32(8):107210.

3. McKevitt C, Fudge N, Redfern J, Sheldenkar A, Crichton S, Rudd AR, Forster A, et al. Self-reported long-term needs after stroke. Stroke. 2011 May;42(5):1398–403.

4. Quinn K, Murray C, Malone C. Spousal experiences of coping with and adapting to caregiving for a partner who has a stroke: a meta-synthesis of qualitative research. Disabil Rehabil. 2014;36(3):185–98.

5. King D, Wittenberg R, Patel A, Quayyum Z, Berdunov V, Knapp M. The future incidence, prevalence and costs of stroke in the UK. Age Ageing. 2020 Feb 27;49(2):277–82.

6. Amarenco P, Steering C, Investigators of the ToP. Five-Year Risk of Stroke after TIA or Minor Ischemic Stroke. N Engl J Med. 2018 Oct 18;379(16):1580–1.

7. Flach C, Muruet W, Wolfe CDA, Bhalla A, Douiri A. Risk and Secondary Prevention of Stroke Recurrence: A Population-Base Cohort Study. Stroke. 2020 Aug;51(8):2435–44.

8. Buckley BJR, Harrison SL, Hill A, Underhill P, Lane DA, Lip GYH. Stroke-Heart Syndrome: Incidence and Clinical Outcomes of Cardiac Complications Following Stroke. Stroke. 2022 May;53(5):1759–63.

9. Saunders DH, Sanderson M, Hayes S, Johnson L, Kramer S, Carter DD, Jarvis H, et al. Physical fitness training for stroke patients. Cochrane Database Syst Rev. 2020 Mar 20;3(3):CD003316.

10. Moncion K, Rodrigues L, Wiley E, Noguchi KS, Negm A, Richardson J, MacDonald MJ, et al. Aerobic exercise interventions for promoting cardiovascular health and mobility after stroke: a systematic review with Bayesian network meta-analysis. Br J Sports Med. 2024 Mar 21;58(7):392–400.

11. English C, Hillier SL, Lynch EA. Circuit class therapy for improving mobility after stroke. Cochrane Database Syst Rev. 2017 Jun 2;6(6):CD007513.

12. Mah SM, Goodwill AM, Seow HC, Teo WP. Evidence of High-Intensity Exercise on Lower Limb Functional Outcomes and Safety in Acute and Subacute Stroke Population: A Systematic Review. Int J Environ Res Public Health. 2022 Dec 22;20(1).

13. MacKay-Lyons M, Billinger SA, Eng JJ, Dromerick A, Giacomantonio N, Hafer-Macko C, Macko R, et al. Aerobic Exercise Recommendations to Optimize Best Practices in Care After Stroke: AEROBICS 2019 Update. Phys Ther. 2020 Jan 23;100(1):149–56.

14. Billinger SA, Arena R, Bernhardt J, Eng JJ, Franklin BA, Johnson CM, MacKay-Lyons M, et al. Physical activity and exercise recommendations for stroke survivors: a statement for healthcare professionals from the American Heart Association/American Stroke Association. Stroke. 2014 Aug;45(8):2532–53.

15. Ambrosetti M, Abreu A, Corra U, Davos CH, Hansen D, Frederix I, Iliou MC, et al. Secondary prevention through comprehensive cardiovascular rehabilitation: From knowledge to implementation. 2020 update. A position paper from the Secondary Prevention and Rehabilitation Section of the European Association of Preventive Cardiology. Eur J Prev Cardiol. 2021 May 14;28(5):460–95.

16. Intercollegiate Stroke Working Party. National Clinical Guideline for Stroke for the UK and Ireland. London: https://www.strokeguideline.org; 2023.

17. Gaskins NJ, Bray E, Hill JE, Doherty PJ, Harrison A, Connell LA. Factors influencing implementation of aerobic exercise after stroke: a systematic review. Disabil Rehabil. 2021 Aug;43(17):2382–96.

18. Connell LA, Klassen TK, Janssen J, Thetford C, Eng JJ. Delivering Intensive Rehabilitation in Stroke: Factors Influencing Implementation. Phys Ther. 2018 Apr 1;98(4):243–50.

19. Regan EW, Handlery R, Stewart JC, Pearson JL, Wilcox S, Fritz S. Feasibility of integrating survivors of stroke into cardiac rehabilitation: A mixed methods pilot study. PLoS One. 2021;16(3):e0247178.

20. Clague-Baker N, Robinson T, Gillies CL, Drewry S, Hagenberg A, Singh S. Adapted cardiac rehabilitation for people with sub-acute, mild-to-moderate stroke: a mixed methods feasibility study. Physiotherapy. 2022 Jun;115:93–101.

21. Heron N, Kee F, Mant J, Cupples ME, Donnelly M. Rehabilitation of patients after transient ischaemic attack or minor stroke: pilot feasibility randomised trial of a home-based prevention programme. Br J Gen Pract. 2019 Oct;69(687):e706–e14.

22. R Core Team. R: A Language and Environment for Statistical Computing. R Foundation for Statistical Computing. Vienna, Austria 2025.

23. Ruff J, Udeh B, Linder S. Cardiac Rehabilitation for Persons with Stroke: A Cost-Effectiveness Analysis. Clin Rehabil. 2025 Feb;39(2):153–60.

24. Jones J, Buckley B, Furze G, Sheppard G, editors. Cardiovascular Prevention and Rehabilitation in Practice. Second Edition ed. Chichester: Wiley-Blackwell; 2020.

25. Yaghi S, Engelter S, Del Brutto VJ, Field TS, Jadhav AP, Kicielinski K, Madsen TE, et al. Treatment and Outcomes of Cervical Artery Dissection in Adults: A Scientific Statement From the American Heart Association. Stroke. 2024 Mar;55(3):e91–e106.

26. Vallance JK, Hale I, Hansen G. Commentary: Physical activity after patent foramen ovale (PFO)-associated stroke: a personal narrative and call to action. Top Stroke Rehabil. 2023 Apr;30(3):304–8.

