## Supplementary material for "Cardiorespiratory training after stroke or transient ischemic attack in the United Kingdom: a national survey": Online Supplementary Materials

### Supplementary Material Table A. **What is the frequency of the supervised sessions?**

Question asked to Groups 1 and 2 (Those who reported integrating CR into routine stroke rehabilitation, and those who reported working in a specialist CR service and provided CR training to those with stroke or TIA)

| Frequency of supervised sessions | Group 1: Integrating CR into stroke rehabilitation | Group 2: CR training service including stroke | p-value |
|---|---|---|---|
| Less than once per week | 25 (26.9%) | 0 (0.0%) | 0.003 |
| Once per week | 54 (58.1%) | 24 (77.4%) | 0.090 |
| More than once per week | 32 (34.4%) | 11 (35.5%) | 1 |

### Supplementary Material Table B. **I am more likely to incorporate cardiorespiratory training with individuals who… (please tick all that apply).**

Question asked to Group 1 only (Those who reported integrating CR into routine stroke rehabilitation)

| Responses | Count (percentage) |
|---|---|
| I incorporate cardiorespiratory training with all patients | 41 (38.0%), n = 108 |
| Individuals with minimal physical disability | 59 (54.6%), n = 108 |
| Individuals with minimal cognitive disability | 57 (52.8%), n = 108 |
| Individuals who were more physically active before their stroke or TIA | 55 (50.9%), n = 108 |
| Individuals with minimal communication disabilities | 44 (40.7%), n = 108 |
| Younger individuals | 43 (39.8%), n = 108 |
| Individuals who were less physically active before their stroke or TIA | 33 (30.6%), n = 108 |
| Individuals who are deemed to be at lower risk of cardiovascular disease | 32 (29.6%), n = 108 |
| Older individuals | 29 (26.9%), n = 108 |
| Individuals who are deemed to be at higher risk of cardiovascular disease | 28 (25.9%), n = 108 |
| Individuals with significant communication disabilities | 24 (22.2%), n = 108 |
| Individuals with significant physical disability | 21 (19.4%), n = 108 |
| Individuals with significant cognitive disability | 9 (8.3%), n = 108 |
| Other | 9 (8.3%), n = 108 |

### Supplementary Material Table C. **How are people who have had a stroke or TIA eligible? Please tick all that apply.**

Question asked to Group 2 only (Those who reported working in a specialist CR service and provided CR training to those with stroke or TIA)

| Responses | Count (percentage) |
|---|---|
| Stroke or TIA diagnosis alone | 4 (10.5%), n = 38 |
| Stroke diagnosis alone | 6 (15.8%), n = 38 |
| TIA diagnosis alone | 1 (2.6%), n = 38 |
| Stroke diagnosis only if they have a secondary diagnosis such as a cardiac condition | 28 (73.7%), n = 38 |
| TIA diagnosis only if they have a secondary diagnosis such as a cardiac condition | 26 (68.4%), n = 38 |

### Supplementary Material Table D. **Are people with stroke eligible regardless of the severity of their stroke impairments and disability? Please tick all that apply.**

Question asked to Group 2 only (Those who reported working in a specialist CR service and provided CR training to those with stroke or TIA)

| Responses | Count (percentage) |
|---|---|
| Stroke or TIA diagnosis alone | 4 (10.5%), n = 38 |
| Stroke diagnosis alone | 6 (15.8%), n = 38 |
| TIA diagnosis alone | 1 (2.6%), n = 38 |
| Stroke diagnosis only if they have a secondary diagnosis such as a cardiac condition | 28 (73.7%), n = 38 |
| TIA diagnosis only if they have a secondary diagnosis such as a cardiac condition | 26 (68.4%), n = 38 |

### Supplementary Material Table E. **Please list any stroke-specific precautions or contraindications to cardiorespiratory training that you use? Please tick all that apply.**

Question asked to Groups 1 and 2 (Those who reported integrating CR into routine stroke rehabilitation, and those who reported working in a specialist CR service and provided CR training to those with stroke or TIA)

| Responses | Count (percentage) |
|---|---|
| Unknown cause of stroke | 43 (37.7%), n = 114 |
| Unknown cause of TIA | 27 (23.7%), n = 114 |
| Patent Foramen Ovale / septal defect | 38 (33.3%), n = 114 |
| Cervical arterial dissection | 58 (50.9%), n = 114 |
| Within two weeks of stroke/TIA | 40 (35.1%), n = 114 |
| Within twelve weeks of stroke/TIA | 7 (6.1%), n = 114 |
| Within other period of time | 3 (2.6%), n = 114 |
| Post-stroke fatigue | 54 (47.4%), n = 114 |

| | |
|---|---|
| Exertional fatigue | 38 (33.3%), n = 114 |
| Cognitive impairment | 56 (49.1%), n = 114 |
| Physical impairment | 44 (38.6%), n = 114 |
| Communication impairment | 26 (22.8%), n = 114 |
| Other - Unknown | 1 (0.9%), n = 114 |
| Other - Tonal changes | 1 (0.9%), n = 114 |
| Other - barriers to engagement | 1 (0.9%), n = 114 |
| Other - awaiting carotid US or 72 hour tape | 1 (0.9%), n = 114 |
| Other - type of stroke (ischaemic vs. haemorrhagic) | 1 (0.9%), n = 114 |

Supplementary Material Table F. **Are these [referring to responses given summarised in Table D] contraindications (i.e. cannot proceed with cardiorespiratory training), or precautions (things to monitor closely or reduce intensity of cardiorespiratory training)? Please indicate for each option.**

| Factor | Precaution | Contraindication |
|---|---|---|
| Unknown cause of stroke | 33 (31.4%) | 10 (20.8%) |
| Unknown cause of TIA | 18 (17.1%) | 9 (18.8%) |
| Patent Foramen Ovale / septal defect | 25 (23.8%) | 11 (22.9%) |
| Cervical arterial dissection | 33 (31.4%) | 23 (47.9%) |
| Within two weeks of stroke/TIA | 22 (21.0%) | 16 (33.3%) |
| Within twelve weeks of stroke/TIA | 6 (5.7%) | 0 (0.0%) |
| Within other period of time | 3 (2.9%) | 0 (0.0%) |
| Post-stroke fatigue | 52 (49.5%) | 1 (2.1%) |
| Exertional fatigue | 37 (35.2%) | 0 (0.0%) |
| Cognitive impairment | 49 (46.7%) | 6 (12.5%) |
| Physical impairment | 41 (39.0%) | 3 (6.2%) |
| Communication impairment | 23 (21.9%) | 3 (6.2%) |
| Other - Unknown | 1 (1.0%) | 0 (0.0%) |
| Other - Tonal changes | 1 (1.0%) | 0 (0.0%) |
| Other - barriers to engagement | 1 (1.0%) | 0 (0.0%) |
| Other - awaiting carotid US or 72 hour tape | 1 (1.0%) | 0 (0.0%) |
| Other - type of stroke (ischaemic vs. haemorrhagic) | 1 (1.0%) | 0 (0.0%) |

Supplementary Material Table G. **What concerns (if any) do you have about incorporating cardiorespiratory training into the rehabilitation of people who have had a stroke or TIA?**

Question asked to all four groups

| Concerns | Group 1: Integrating CR into stroke rehabilitation (n = 113) | Group 2: CR training service including stroke (n = 40) | Group 3: Not integrating CR into stroke rehabilitation (n = 74) | Group 4: CR training service not including stroke (n = 26) | P-value | Significant (p<0.05) between group differences |
|---|---|---|---|---|---|---|
| No concerns | 9 (8.0%, n = 113) | 8 (20.0%, n = 40) | 12 (16.2%, n = 74) | 6 (23.1%, n = 26) | 0.0576 | None |
| Time may be better spent focussing on traditional stroke rehabilitation | 19 (16.8%, n = 113) | 6 (15.0%, n = 40) | 19 (25.7%, n = 74) | 4 (15.4%, n = 26) | 0.412 | None |
| Risk of serious adverse event whilst exercising (e.g. cardiovascular event) | 54 (47.8%, n = 113) | 7 (17.5%, n = 40) | 30 (40.5%, n = 74) | 2 (7.7%, n = 26) | <0.001 | 1 vs. 2<br>1 vs. 4<br>3 vs. 4 |
| Risk of falling | 33 (29.2%, n = 113) | 13 (32.5%, n = 40) | 11 (14.9%, n = 74) | 7 (26.9%, n = 26) | 0.0814 | None |

| | | | | | | |
|---|---|---|---|---|---|---|
| Increased fatigue | 33 (29.2%, n = 113) | 7 (17.5%, n = 40) | 22 (29.7%, n = 74) | 8 (30.8%, n = 26) | 0.476 | None |
| Increase in compensatory/abnormal movement patterns | 36 (31.9%, n = 113) | 7 (17.5%, n = 40) | 19 (25.7%, n = 74) | 5 (19.2%, n = 26) | 0.28 | None |
| Fear of liability | 13 (11.5%, n = 113) | 3 (7.5%, n = 40) | 8 (10.8%, n = 74) | 1 (3.8%, n = 26) | 0.742 | None |
| Other - ability to achieve high intensity with people with severe stroke impairments | 3 (2.7%, n = 113) | 0 (0.0%, n = 40) | 1 (1.4%, n = 74) | 0 (0.0%, n = 26) | 0.888 | None |
| Other - not considered safe by medical team | 0 (0.0%, n = 113) | 0 (0.0%, n = 40) | 1 (1.4%, n = 74) | 0 (0.0%, n = 26) | 0.553 | None |
| Other - resources (including monitoring equipment, exercise equipment, space, time, and staff) | 9 (8.0%, n = 113) | 2 (5.0%, n = 40) | 4 (5.4%, n = 74) | 3 (11.5%, n = 26) | 0.682 | None |
| Other - new to guidelines and not traditional practice in setting | 0 (0.0%, n = 113) | 0 (0.0%, n = 40) | 1 (1.4%, n = 74) | 0 (0.0%, n = 26) | 0.553 | None |
| Other - increase waiting list for cardiac patients | 0 (0.0%, n = 113) | 0 (0.0%, n = 40) | 0 (0.0%, n = 74) | 1 (3.8%, n = 26) | 0.103 | None |
| Other - lack of training for staff | 0 (0.0%, n = 113) | 1 (2.5%, n = 40) | 1 (1.4%, n = 74) | 0 (0.0%, n = 26) | 0.397 | None |
| Other - knowledge gaps | 1 (0.9%, n = 113) | 0 (0.0%, n = 40) | 1 (1.4%, n = 74) | 1 (3.8%, n = 26) | 0.402 | None |
| Other - not aligned with patient goals | 0 (0.0%, n = 113) | 0 (0.0%, n = 40) | 1 (1.4%, n = 74) | 0 (0.0%, n = 26) | 0.553 | None |
| Other - not appropriate due to physical disabilities | 0 (0.0%, n = 113) | 0 (0.0%, n = 40) | 1 (1.4%, n = 74) | 0 (0.0%, n = 26) | 0.553 | None |

### Supplementary Material Table H. **What is currently preventing you from delivering cardiorespiratory training into the rehabilitation of people who have had a stroke or TIA?**

Question asked to Groups 3 and 4 (Those who reported not integrating CR into routine stroke rehabilitation, and those who reported working in a specialist CR service and did not provide (or rarely provided) CR training to those with stroke or TIA).

| **Barriers to delivering CR training** | **Group 3: Not integrating CR into stroke rehabilitation (n = 74)** | **Group 4: CR training service not including stroke (n = 26)** | **P-value** |
|---|---|---|---|
| Commissioning - Service is not commissioned for people who have had a stroke | 18 (24.3%), n = 74 | 16 (61.5%), n = 26 | 0.001 |
| Commissioning - Service is not commissioned for people who have had a TIA | 15 (20.3%), n = 74 | 15 (57.7%), n = 26 | <0.001 |
| Resources - Inadequate staffing | 33 (44.6%), n = 74 | 10 (38.5%), n = 26 | 0.754 |

| | | | |
|---|---|---|---|
| Resources - Inadequate space or environment | 33 (44.6%), n = 74 | 4 (15.4%), n = 26 | 0.016 |
| Resources - Inadequate equipment | 30 (40.5%), n = 74 | 3 (11.5%), n = 26 | 0.014 |
| Resources - Lack of resources for screening or risk-assessment | 26 (35.1%), n = 74 | 6 (23.1%), n = 26 | 0.374 |
| Resources - Medical team unable to support screening or risk-assessments | 21 (28.4%), n = 74 | 3 (11.5%), n = 26 | 0.144 |
| Skills/knowledge regarding cardiorespiratory training | 54 (73.0%), n = 74 | 2 (7.7%), n = 26 | <0.001 |
| Skills/knowledge regarding stroke | 3 (4.1%), n = 74 | 8 (30.8%), n = 26 | <0.001 |
| Lack of guidelines regarding screening or risk-assessment | 30 (40.5%), n = 74 | 10 (38.5%), n = 26 | 1 |
| People who have had a stroke or TIA are not referred to our service | 0 (0.0%), n = 74 | 14 (53.8%), n = 26 | <0.001 |
| Patient cohort not suitable for cardiorespiratory training | 5 (6.8%), n = 74 | 3 (11.5%), n = 26 | 0.425 |
| Other - clinical need | 1 (1.4%), n = 74 | 0 (0.0%), n = 26 | 1 |
| Other - currently planning to | 1 (1.4%), n = 74 | 0 (0.0%), n = 26 | 1 |
| Other - appropriateness of caseload | 5 (6.8%), n = 74 | 0 (0.0%), n = 26 | 0.323 |
| Other - not sure | 0 (0.0%), n = 74 | 1 (3.8%), n = 26 | 0.260 |
| Other - focus on functional rehab | 1 (1.4%), n = 74 | 0 (0.0%), n = 26 | 1 |
| Other - not referred | 1 (1.4%), n = 74 | 0 (0.0%), n = 26 | 1 |
| Other - requires a new pathway | 1 (1.4%), n = 74 | 0 (0.0%), n = 26 | 1 |

### Survey

#### Eligibility Questions/Confirmation

- I am a physiotherapist or exercise professional working in the United Kingdom.

**[If a participant does not confirm this, they will be sent an automated ineligibility message (see below).]**

- My role includes providing rehabilitation to people who have had a stroke or TIA.
- My role includes providing or supporting the delivery of cardiac rehabilitation or a similar service (this option does not need to include working with people who have had a stroke or TIA).
- Neither of the above.

[If a participant answers the final option from the list above, they will be sent an automated ineligibility message, which will read:]

Thank you for your time and interest in this project. From your answer to the previous question, it looks like you may not be eligible to take part. To take part you must be a physiotherapist or exercise professional practising in the United Kingdom and working in stroke rehabilitation or in cardiac rehabilitation services. If you think there has been a mistake, please get in touch with Dr Peter Hartley for further assistance.

[Section 1 – Filter Questions]

*To ask you the most appropriate questions, this first section is designed to understand the type of service that you work in.*

**1.1.** Which sector(s) do you work in. Please select all that apply.

Response options (multiple choice – can select multiple):

- National Health Service
- Private practice
- Charitable
- Local Authority Services
- Other (please specify)

**[If selected more than one option in Q1.1. go to question 1.2. if only one option go straight to 1.3]**

**1.2. [If selected more than one option in Q1.1.] To help us interpret the survey, please base your answers on one sector only. Select the most relevant option (e.g. the sector most related to cardiorespiratory training after stroke or TIA, or where you work the most hours).**

Response options (single choice):

- National Health Service
- Private practice
- Charitable
- Local Authority Services
- Other (please specify)

**2.1. As part of your clinical practice do you ever incorporate cardiorespiratory training for people after stroke or TIA as part of routine rehabilitation or as part of a specialist service?**

Response options (multiple choice – can select only one):

- Yes (includes people seen for other reasons, but who have a history of stroke/TIA) [goes to question 2]
- No, or very rarely [goes to question 5]

***Participants who responded to question 2 with 'Yes'***

**2.2. Please select the box that best describes how you deliver cardiorespiratory training to people who have had a stroke or TIA.**

Response options (multiple choice – can select only one):

- Specialist service that delivers cardiorespiratory training (e.g. Cardiac Rehabilitation, or secondary prevention services that include people who have had a stroke/TIA, or are specifically adapted to or designed for this population) ***[Leads to question 3, skips question 4].***
- Cardio-respiratory training incorporated into routine stroke rehabilitation (e.g. stroke inpatient, outpatient, or community rehabilitation services that provide cardiorespiratory training as part of routine rehabilitation to some or all patients) ***[Leads to question 4, skips question 3].***

**[Section 3 – aimed at people delivering CR type services]**

***Participants who responded to question 2 with '***Specialist service' or 'Both'

***About your service:*** *in this section we'd like to know more about the service that you work in, and how cardiorespiratory is delivered to people who have had a stroke or TIA.*

**3.1. What type of service do you work in? Select one option only, if you work in multiple services, please select the most relevant option to delivering cardiorespiratory training to people who have had a stroke or TIA.**

Single choice (can choose one option only)

- Inpatient cardiology wards
- Inpatient respiratory wards
- Cardiac rehabilitation (i.e. outpatient/community-based Phase II to IV service)
- Other outpatient/community cardiac service
- Outpatient/community respiratory service
- Other (please specify)

#### 3.2. How are patients referred to the service? Please tick all that apply.

Multiple choice (can choose multiple options)

- Self-referral
- Referrals from secondary or tertiary care (e.g. hospital consultant/services)
- Referrals from primary care (e.g. GPs, community teams)
- Other (please specify)

#### 3.3. How are people who have had a stroke or TIA eligible? Please tick all that apply.

Multiple choice (can choose multiple options)

- Stroke diagnosis alone.
- Stroke diagnosis only if they have a secondary diagnosis such as a cardiac condition.
- TIA diagnosis alone.
- TIA diagnosis only if they have a secondary diagnosis such as a cardiac condition.
- Other (please specify)

#### 3.4. Are people with stroke eligible regardless of the severity of their stroke impairments and disability? Please tick all that apply.

Multiple choice (can choose multiple options)

- No eligibility criteria that may relate to the severity of stroke impairments or disability
- Individuals must be able to walk unaided
- Individuals must be able to exercise safely unsupervised
- Eligibility criteria based on physical impairments or ability (please specify)
- Eligibility criteria based on communication impairments (please specify)
- Eligibility criteria based on cognitive impairments (please specify)
- Other (please specify)

#### 3.5. What proportion of the service users do you estimate have a diagnosis of stroke or TIA?

Two separate numeric responses for stroke and TIA

#### 3.6. What is the typical time since the patients have had a stroke after which you would start incorporating cardiorespiratory training into their rehabilitation? Please tick all that apply.

Multiple choice (can choose one option)

- 0–2 weeks since stroke
- 3–12 weeks since stroke
- 13+ weeks since stroke
- Other (please specify)

Free text response:

#### 3.7. Do you have a formal screening tool or process (e.g. medical review, ECG) to follow before you incorporate cardiorespiratory training into a patient's rehabilitation? Please tick all that apply.

Multiple choice (can choose multiple options)

- Doctor assessment following set process (e.g. checklist)
- Doctor assessment with no set process
- Other healthcare professional review following set process/checklist (please specify which healthcare professional)
- Other healthcare professional review not following set process (please specify which healthcare professional)
- Patient self-reported questionnaire
- Screening process routinely includes ECG
- Screening process routinely includes cardiopulmonary exercise testing (CPET)
- Screening process routinely includes sub-maximal exercise test
- Screening process routinely includes a field-based assessment e.g. (6-min walk test, cycle-ergometer test)
- Other (please specify)

#### 3.8. Please list any stroke-specific precautions or contraindications to cardiorespiratory training that you use? Please tick all that apply.

Multiple choice (if ticked, expand to describe)

- Unknown cause of stroke
- Unknown cause of TIA
- Patent Foramen Ovale (or other atrial and ventricular septal defects) (please expand)
- Cervical arterial dissection
- Within two weeks of stroke/TIA
- Within twelve weeks of stroke/TIA
- Within other period of time since stroke/TIA
- Post-stroke fatigue
- Exertional fatigue
- Cognitive impairment
- Physical impairment
- Communication impairments (e.g. expressive dysphasia)
- Other (please specify) [Free text response:]

**[Could this pipe through to another question where they have to indicate whether the options they selected are precautions or contraindications. This should also have a free text box at end 'Please add any comments if you wish to expand']**

**3.8.1. Are these contraindications (i.e. cannot proceed with cardiorespiratory training), or precautions (things to monitor closely or reduce intensity of cardiorespiratory training)? Please indicate for each option.**

**3.9. Do you set intensity targets for your patients, and if so, how? Please tick all that apply.**

Multiple choice (can choose multiple options)

- Do not set intensity targets
- Maximal cardiopulmonary exercise testing (CPET)
- Submaximal cardiopulmonary exercise testing (CPET)
- Six-minute walk test
- Cycle ergometer test
- Shuttle walk test
- Step test

- Other (please specify)

#### 3.10. Do you measure intensity of training, if so how? Please tick all that apply.

Multiple choice (can choose multiple options)

- Not measured
- Heart-rate monitor
- Patient self-report using Borg Breathlessness Scale
- Patient self-report using another tool (please specify)
- Clinician's judgement
- Other (please specify)

#### 3.11. What is the format of the supervised sessions? Please tick all that apply.

Multiple choice (can choose multiple options)

- Group in-person
- 1:1 in-person
- Group virtual
- 1:1 virtual
- Other (please specify)

#### 3.12. What is the frequency of the supervised sessions? Please tick all that apply.

Multiple choice (can choose multiple options)

- Weekly
- Less than once a week
- More than once a week

#### 3.13. Where is the location of the supervised sessions? Please tick all that apply.

Multiple choice (can choose multiple options)

- In-hospital - inpatients

- In-hospital - outpatients
- Participant's home
- GP practice
- Private gym
- Private rehabilitation centre
- Community centre
- Leisure centre
- Premises of charity
- Other (please specify)

#### 3.14. What is the typical duration in weeks of the supervised sessions?

[Numeric response]

Other (please specify)

#### 3.15. Do staff complete any training or competencies to deliver the cardiorespiratory training? Please tick all that apply.

Multiple choice (can choose multiple options)

- No competencies
- No training
- Formal competencies not specific to stroke/TIA
- Formal competencies specific to stroke/TIA
- Informal process to check competence not specific to stroke/TIA
- Informal process to check competence specific to stroke/TIA
- Formal training not specific to stroke
- Formal training specific to stroke
- Informal training not specific to stroke
- Informal training specific to stroke
- Other (please specify)

***Challenges and risks:*** *in this section we'd like to find out more about your opinions of the challenges and risks of cardiorespiratory training after stroke or TIA.*

**3.16. What concerns (if any) do you have about incorporating cardiorespiratory training into the rehabilitation of people who have had a stroke or TIA?**

Multiple choice (can choose multiple options)

- No concerns
- Time may be better spent focussing on traditional stroke rehabilitation
- Risk of serious adverse event whilst exercising (e.g. cardiovascular event)
- Risk of falling
- Increased fatigue
- Increase in compensatory/abnormal movement patterns
- Fear of liability
- Other (please specify).

**3.17. Please rate your current confidence to deliver cardiorespiratory training to people with stroke/TIA regardless of level of impairment on the scale below**

[A scale from 'Fully confident' to 'Not at all confident']

**3.18. If not fully confident, what would increase your confidence?**

Free text response

**[Section 3 – aimed at people delivering stroke rehabilitation]**

***About your service:*** *in this section we'd like to know more about the service that you work in, and how cardiorespiratory is delivered to people who have had a stroke or TIA.*

***Participants who responded to question 2 with*** **'Incorporated into routine stroke-rehabilitation'**

**4.1. [Same wording as Q3.1. but different options]**

Multiple choice (can choose multiple options)

- Acute stroke unit
- Inpatient rehabilitation unit – specialist
- Inpatient rehabilitation unit – non specialist
- Outpatient/community neuro-rehabilitation service
- Reablement or Integrated Care team or other community rehabilitation team linked with short-term care service
- Other (please specify)

Free text response:

#### 4.2. I am more likely to incorporate cardiorespiratory training with individuals who… (please tick all that apply)

Multiple choice (can choose multiple options)

- have minimal physical disability
- have significant physical disability
- have minimal cognitive disability
- have significant cognitive disability
- have minimal communication disabilities
- have significant communication disabilities
- were more physically active before their stroke or TIA
- were less physically active before their stroke or TIA
- are deemed to be at lower cardiovascular risk
- are deemed to be at higher cardiovascular risk
- are younger
- are older
- I incorporate cardiorespiratory training with all patients
- Other (please specify)

#### 4.3. [Same wording as 3.5. What is the typical time since the patients have had a stroke after which you would start incorporating cardiorespiratory training into their rehabilitation?]

**4.4.** **[Same wording as 3.6. - Do you have a formal screening tool or process (e.g. medical review, ECG) to follow before you incorporate cardiorespiratory training into a patient's rehabilitation?]**

**4.5.** **[Same wording as 3.7. - Could you list any stroke-specific precautions or contraindications to cardiorespiratory training, and how they impact your management?]**

**4.6.** **[Same wording as 3.8. Do you set intensity targets for your patients? If so, do you use exercise testing (e.g. CPET, submaximal testing, 6 min walk test), clinical judgement, or another method to set the targets?**

**4.7.** **[Same wording as 3.9. Do you measure intensity of training, if so how?]**

**4.8.** **[Same wording as 3.10. What is the format of the supervised sessions?**

**4.9.** **[Same wording as 3.11. What is the frequency of the supervised sessions?]**

**4.10.** **[Same wording as 3.12. - Where is the location of the supervised sessions?]**

**4.11.** **[Same wording as 3.13 - What is the typical duration in weeks of the supervised sessions?]**

**4.12.** **[Same wording as 3.14. - Do staff complete any training or competencies to deliver the cardiorespiratory training?]**

***Challenges and risks: in this section we'd like to find out more about your opinions of the challenges and risks of cardiorespiratory training after stroke or TIA.***

**4.13.** **[Same wording as 3.16. - What concerns (if any) do you have about incorporating cardiorespiratory training into the rehabilitation of people who have had a stroke or TIA?]**

**4.14.** **[Same wording as 3.17] Please rate your current confidence to deliver cardiorespiratory training to people with stroke/TIA regardless of level of impairment on the scale below**

**4.15. [Same wording as 3.18] If not fully confident, what would increase your confidence?**

***Participants who responded to question 2 with 'No'***

***About your service: in this section we'd like to know more about the service that you work in.***

5.1. **[Same wording as Q3.1. but different options]**

Response options (multiple choice – can select multiple):

- Acute stroke unit
- Inpatient rehabilitation unit – specialist
- Inpatient rehabilitation unit – non specialist
- Inpatient cardiology wards
- Inpatient respiratory wards
- Cardiac rehabilitation (i.e. outpatient/community-based Phase II to IV service)
- Other outpatient/community cardiac service
- Stroke early supported discharge service
- Outpatient/community respiratory service
- Outpatient/community neuro-rehabilitation service
- Reablement or Integrated Care team or other community rehabilitation team linked with short-term care service
- Other (please specify)

***Understanding challenges and risks:*** *in this section we'd like to find out more about your opinions of the challenges and risks of cardiorespiratory training after stroke or TIA.*

**5.2. What is currently preventing you from delivering cardiorespiratory training into the rehabilitation of people who have had a stroke or TIA?**

Multiple choice (can choose multiple options)

Commissioning

- Service is not commissioned for people who have had a stroke
- Service is not commissioned for people who have had a TIA

Resources

- Inadequate staffing
- Inadequate space or environment
- Inadequate equipment
- Lack of resources for screening or risk-assessment
- Medical team unable to support screening or risk-assessments
- Other inadequate resources [pipe through to text entry question and ask about each individually]

Skills and knowledge and guidance

- Skills/knowledge regarding cardiorespiratory training
- Skills/knowledge regarding stroke
- Staff require other skills or knowledge [pipe through to text entry question and ask about each individually]
- Lack of guidelines regarding screening or risk-assessment

Other

- People who have had a stroke or TIA are not referred to our service
- Patient cohort not suitable for cardiorespiratory training
- Other (please specify)

**5.3. [Same wording as 3.16. and 4.13. - What concerns (if any) do you have about incorporating cardiorespiratory training into the rehabilitation of people who have had a stroke or TIA?**

**5.4. Do you have (or know of) plans to provide cardiorespiratory training to people who have had a stroke or TIA?**

Response options (single choice):

- Yes, for both TIA and stroke [Got to 5.5.]

- Yes, for TIA only [Go to 5.5.]
- Yes, for stroke only [Go to 5.5.]
- No [Go to 5.6.]

***Participants who responded to question 5.4 with 'Yes'***

#### 5.5. Please could you provide details of these plans, and what the service or rehabilitation might look like?

Free text response:

***Referrals:*** *In this section we'd like to find out more about services you are able to refer people to for cardiorespiratory training after stroke or TIA.*

#### 5.6. Are you able to refer people who have had a stroke or TIA to a service that delivers cardiorespiratory training?

Response options (single choice):

- Yes, both TIA and stroke
- Yes, TIA only
- Yes, stroke only
- No **[Go to question 6]**

**[Participants who responded to question 5.5. with 'Yes']**

#### 5.7. What services do you refer to?

Multiple choice (can choose multiple options)

- NHS/publicly funded cardiac rehabilitation service
- Charity funded cardiac rehabilitation service
- NHS/publicly funded stroke or TIA specific outpatient cardiorespiratory training service
- Charity funded stroke or TIA specific outpatient cardiorespiratory training service
- NHS/publicly funded cardiorespiratory training service (not cardiac rehabilitation)
- Charity funded cardiorespiratory training service (not cardiac rehabilitation)
- Other (please specify)

#### 5.8. How you decide who to refer? Please select all that apply.

Multiple choice (can choose multiple options)

- Every individual who has had a stroke are offered referral
- Every individual who has had a TIA are offered referral
- Individuals with cardiac conditions are offered referral
- Individuals able to walk unaided are offered referral
- Individuals who are deemed safe to exercise unsupervised are offered referral
- Other criteria based on physical impairments or ability (please specify)
- Other criteria based on communication impairments (please specify)
- Other criteria based on cognitive impairments (please specify)
- Other (please specify)

#### 5.9. Do you do any pre-screening? Please select all that apply.

Multiple choice (can choose multiple options)

- No pre-screening
- Physician assessment following set process (e.g. checklist)
- Physician assessment with **no** set process
- Other healthcare professional review following set process/checklist
- Other healthcare professional review **not** following set process/checklist
- Patient self-reported questionnaire
- Screening process routinely includes ECG
- Other (please specify)

### [Section 4 – Any other information]

#### 6.1. Do you have any other comments you would like to make, including what you feel an ideal service model that provides cardiorespiratory training to people after stroke or TIA should look like?

Free text response:

### [Section 5 - Demographic information]

***Thank you so much for all your answers so far. Before you go, we would like to learn a little bit more about you and your role. You are free to skip any of these questions.***

#### 7.1. What is your job role?

- Physiotherapist
- Clinical exercise physiologist
- Other exercise professional (please specify)

#### 7.2. Which geographical area do you practice in?

Response options (multiple choice – can select only one)

- Scotland
- Wales
- Northern Ireland
- England – North East
- England – North West
- England – Yorkshire and the Humber
- England – East Midlands
- England – West Midlands
- England – South East
- England – East of England
- England – South West
- England – London

**7.3. Which NHS Trust, Health Board or organisation do you work for? We will not report results for separate organisations, but will use this to recognise multiple responses from the same organisation. (If self-employed please write 'self-employed' in the box below.)**

Free text response:

**7.4. How many years of professional practice do you have? Please enter a value to the nearest approximate year.**

Response options (enter numeral):

***Thank you for taking part***

**8.1. Are you happy for Thiscovery and the project team to stay in touch? Please select all that apply.**

- I'm happy to be contacted about this project
- I'm happy to be contacted about other, relevant Thiscovery opportunities

Would you like a certificate of participation for Continuing Professional Development (CPD) evidence?

- Yes, I would like a certificate of participation for CPD

The project team at the University of Cambridge will be using the results of this survey to support follow-up research involving virtual interviews. Would you like to be invited to take part in these virtual interviews?

- I'm happy to be contacted by the project team

[If no options selected, survey ends with message:] ***Thank you for your time***

**8.2. Please leave your name and email address, so we can get in touch.**

- First name [free text option]
- Surname [free text option]
- Email address [free text option]

By providing this information you consent to Thiscovery and the project team processing it to update you in line with your preferences. How we manage your data [links to data management document].

**8.3. [Final page to confirm contact details where respondents will be asked to confirm data entered in 8.2. is correct]**

***Thank you for your time***
